# Genome-wide association study of longitudinal urinary albumin excretion in patients with type 1 diabetes

**DOI:** 10.1101/2022.12.19.22283443

**Authors:** Anna M Hutchinson, Wei-Min Chen, Suna Onengut-Gumuscu, Paul Benitez-Aguirre, Fergus J Cameron, Scott T Chiesa, Jennifer J Couper, Maria E Craig, Neil R. Dalton, Denis Daneman, Elizabeth A Davis, John E Deanfield, Kim C Donaghue, Timothy W Jones, Farid H Mahmud, Sally M Marshall, Andrew Neil, Stephen S Rich, M. Loredana Marcovecchio, Chris Wallace

## Abstract

Identifying genetic determinants for longitudinal changes in albumin excretion in individuals with type 1 diabetes may help identify those that are predisposed to renal, retinal and cardiovascular complications. Most studies have focussed on genetic predisposition to diabetic kidney disease and used cross-sectional measurements of urinary albumin excretion, but with limited success. Here, we utilise the wealth of longitudinal data and bio-samples collected from cohorts of childhood-onset type 1 diabetes followed over the last 30 years to describe a novel trajectory phenotype quantifying urinary albumin excretion changes during childhood and adolescence. We conducted a genome-wide association study and fine-mapping analysis for albumin excretion in 1584 individuals, finding one signal for cross-sectional albumin excretion close to *GALNTL6* (rs150766792), which validated in a previous independent study, and a novel genome-wide significant signal for albumin excretion trajectory in the *CDH18* gene region (rs145715205). Our trajectory phenotype quantifies albumin progression and offers a complementary measure to an albumin excretion phenotype based on a single measurement (i.e. most recent data collection) or an average of repeated measurements in longitudinal studies. It can be used to identify genetic or other risk factors which predict better or worse prognosis, thus facilitating the development of new preventive and therapeutic approaches.

## Introduction

Diabetic kidney disease (DKD), the leading cause of end-stage renal disease for people with type 1 diabetes, remains a major determinant of morbidity and cardiovascular mortality (1,2). DKD is characterised by progressive increases in urinary albumin excretion and decline in renal function, which follow different patterns, and occur at variable rates, over the lifetime of individuals with diabetes (1).

Extensive evidence indicates that increases in urinary albumin excretion, even within the normal range, predict renal, retinal and cardiovascular disease risk in adults and youth with type 1 diabetes (3–5). Although hyperglycemia is an important modifiable risk factor for increased albumin excretion, some patients still develop DKD irrespective of glycemic control, suggesting that other factors, either environmental or genetic, are implicated in the pathogenesis of this complication (1,6).

The identification of genetic determinants for albumin excretion is clinically important as it would enable the identification of individuals with type 1 diabetes that are predisposed to vascular complications, and potentially guide the development of new preventive and therapeutic approaches. This is particularly relevant to adolescents with type 1 diabetes, who have a higher risk of complications than adults due to longer diabetes exposure and often have suboptimal glycemic control (7).

To date, genome-wide association studies (GWAS) have only identified a small number of genetic variants that associate with DKD, or more specifically, urinary albumin excretion in individuals with type 1 diabetes (8–14); few studies have included cohorts of youth with type 1 diabetes, and most of the findings have not been replicated (6). This could be due to a combination of relatively small sample sizes, differences in the study populations along with discrepancies across studies in the phenotype definition, ranging from microalbuminuria to end-stage renal disease (6).

Most genetic association studies for albumin-related phenotypes are based on cross-sectional measurements of albumin excretion (9,12–14), but cross-sectional analyses do not account for within-sample variability in albumin measurements over time (15). This is particularly true for youth with type 1 diabetes, who often show early increases in albumin excretion a few years after the diagnosis of diabetes with progression towards abnormal levels particularly during puberty, and potential regression into the normal range at the end of puberty (7,16). An alternative approach would be to investigate longitudinal changes in albumin excretion at an early stage in the development of DKD as a continuous trait, which would require summarising the longitudinal data in a manner suitable for GWAS. This is a challenging task due to the variability of duration and frequency of follow-up in many longitudinal studies.

Utilising the wealth of longitudinal data and bio-samples collected from carefully characterised childhood-onset type 1 diabetes cohorts prospectively followed over the last 30 years, we aimed to derive a novel quantitative phenotype of urinary albumin excretion changes during childhood and adolescence, and investigate novel GWAS genetic associations on this quantitative phenotype.

## Research Design and Methods

### Study Population

The study population consisted of young people with type 1 diabetes recruited into three longitudinal studies: the Oxford Regional Prospective Study (ORPS) (7), the Nephropathy Family Study (NFS) (17) and the Adolescents Type 1 diabetes cardiorenal Intervention Trial (AdDIT) (18) (Table 1). For all three studies, ethical approval was obtained from the local ethics committees. Parents of participants provided written informed consent, and study participants were asked to provide their assent, until they reached an age when they could consent.

**Table 1:**
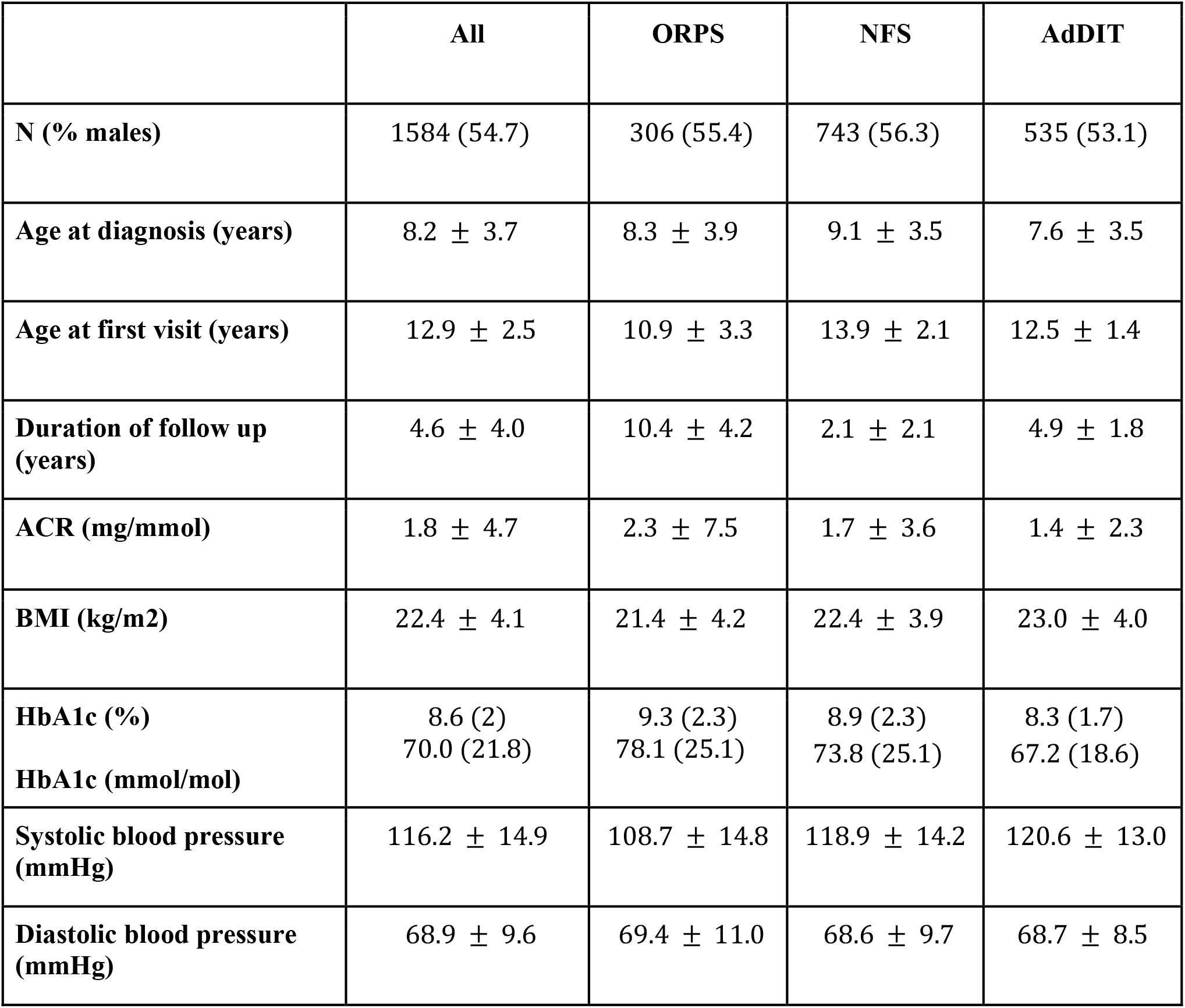
Baseline clinical characteristics for the subjects included in our analysis. Values are expressed as mean±SD for normally distributed continuous variables, median (IQR) for non-normally distributed variables and percentage for categorical variables.

Full details of these cohorts have been previously reported (4,7,19,20). Briefly, the ORPS cohort was established in 1986, and approximately 500 children and adolescents diagnosed with type 1 diabetes when 16 years or younger were recruited between 1986-1996 in a defined geographical area (the Oxford Health Authority) and followed for up to 20 years. Annual assessments included: collection of clinical data, blood samples for HbA1c, and three consecutive urinary samples for the measurement of albumin-creatinine ratios (ACR). At baseline DNA samples were collected from a subgroup of the cohort (*n* = *306*).

The NFS cohort was established between 2000 and 2005, when approximately 1000 young people (10-18 years), who had developed type 1 diabetes before the age of 16 years, were recruited in different UK centres (East Anglia, Birmingham, Bristol and Oxford regions). This cohort has been followed-up with annual assessments of height, weight, BMI, blood pressure, HbA1c and collection of three consecutive early-morning urine specimens for determination of the ACR. At baseline samples for DNA extraction were also collected. The AdDIT cohort included 847 adolescents with type 1 diabetes recruited between the years 2009 and 2013, across three countries (UK, Canada and Australia). In brief, 443 with an ACR in the upper tertile of the normal range (high-ACR group) were recruited to a randomised placebo-controlled trial of ACE-inhibitors and statins, and 404 with an ACR in the lower or middle tertile (low-ACR group) to a parallel observational study. Both groups underwent similar baseline and follow up assessments for a median of 3.9 years, based on a standardised protocol (4,19). For the present study, the study population consisted of 535 adolescents, representing a subgroup of those recruited into the AdDIT Trial and parallel Observational study, with available DNA samples.

### Albumin Excretion

All cohorts were monitored with annual ACR assessments. Urine samples were assessed centrally in a single reference laboratory as previously described (4). We excluded data where less than two consecutive measurements from a single visit were available (224 visits removed from a total of 11,601 visits), or where the ACR measurements from the same visit were highly variable (*standard error of the mean* > *30 mg*/*mmol*; 19 data points removed), as this may indicate measurement errors. We then calculated the geometric mean of the consecutive ACR measurements.

### GWAS Genotyping, Quality Control, and Imputation

We received genotype data for subjects in the ORPS, NFS and AdDIT cohorts. Study participants were genotyped in non-overlapping groups that were not cohort specific, and so we refer to these genotyping groups as groups 1, 2 and 3. Quality control (QC) and imputation were performed separately within each genotyping group (<u>Supplementary Methods</u>; Supplementary Figs 1-5).

Participants in groups 1 and 2 underwent genotyping and QC centrally at the University of Virginia, which has been described previously (11). Samples were genotyped on the HumanCore BeadChip array (Illumina, San Diego, CA), which contains approximately 250,000 genome-wide tag SNPs and 200,000 exome-focused variants. We included an additional QC step to remove 261 chromosome X SNPs that were labelled as heterozygous in males but did not reside in the pseudoautosomal region of chromosome X (Supplementary Table 1). For samples in group 3 which had not been quality controlled, we filtered for low-quality variants (e.g. high missingness, low minor allele frequency and excessive deviation from Hardy–Weinberg equilibrium) and samples (e.g. high missingness, sex mismatch, evidence of relatedness and extreme heterozygosity) (Supplementary Table 2). For samples with replicate data available, we selected the replicate with the best SNP coverage.

We used the Michigan Imputation Server (21) to impute missing genotypes for samples in each genotyping group separately, using the Haplotype Reference Consortium (HRC) panel of 64,976 human haplotypes at 39,235,157 SNPs as the reference data (version r1.1 2016) (22). We specified that the samples were of European ancestry and that any variants with low imputation quality (*R*^*2*^ < *0*.*3*) should be discarded. Following imputation, we removed samples with extreme proportions of homozygous variants and removed any monomorphic SNPs. Across the three genotyping groups, the final imputed and quality controlled data consisted of 11,483,371 SNPs in 2205 samples, for which 1584 samples had phenotype data available and were used in our analysis.

### Multiple Imputation

There was ∼15% missingness for HbA1c, blood pressure and BMI measurements across the study samples, which we accommodated for using a multiple imputation approach (23). Briefly, the multiple imputation pipeline consists of three stages: (i) imputation phase (ii) analysis phase and (iii) pooling phase. We used the mice (“Multivariate Imputation by Chained Equations”) R package (24) to impute 5 complete data sets, using a multilevel modelling approach with subject ID as the class variable and 10 iterations for each imputation, which allowed for sufficient mixing of chains (Supplementary Fig. 6). By specifying a correlation coefficient *r* > *0*.*1* between variables for prediction, we selected ACR, treatment allocation and cohort for the prediction of missing HbA1c values, we selected sex, age, duration of diabetes, HbA1c, treatment allocation and cohort for the prediction of missing BMI values, we selected sex, age, duration of diabetes, HbA1c, treatment allocation (for the AdDIT cohort) and cohort for the prediction of missing systolic blood pressure (SBP) values and we selected ACR, age, duration of diabetes, HbA1c, treatment allocation and cohort for the prediction of missing diastolic blood pressure (DBP) values. To ensure that the imputed values were on the same scale as the observed values (e.g. *HbA1c* > *0*), we used the squeeze function in the mice R package to transform the imputed data values to be within the range of observed data values.

In the analysis phase of multiple imputation, the analysis is implemented separately for each imputed data set. That is, sample-specific phenotypes were derived from each of the 5 imputed data sets (see “Phenotype Definitions” subsection) and 5 separate GWAS were conducted using these phenotype values (see “GWAS Analysis” subsection), resulting in 5 sets of GWAS summary statistics. In the final pooling phase, Rubin’s rules (25) were used to pool the statistics, ultimately deriving a final set of pooled effect sizes, standard errors and p-values (Supplementary Methods).

### Phenotype Definitions

To derive a single albumin excretion phenotype summarising multiple longitudinal ACR measurements (Supplementary Fig. 7), we first used a linear regression model and selected the covariates for inclusion based on previous analyses in these cohorts (7,20). Specifically, the geometric mean of consecutive ACR measurements were logarithmically transformed and used as the dependent variable, whilst sex, duration of diabetes, age, HbA1c, treatment allocation, cohort, BMI, SBP and DBP were selected as covariates in the model. For study samples in ORPS and NFS (where no treatments were allocated) the treatment allocation group was set to the control group in the AdDIT cohort. We extracted the residuals (observed - fitted values) from the model, which correspond to covariate-adjusted ACR measurements.

We first defined a phenotype based on cross-sectional albumin excretion measurements (9,12–14) by selecting the covariate-adjusted ACR measurement from the most recent visit for each subject to derive the “latest ACR phenotype”. Second, we averaged the covariate-adjusted ACR measurements for each subject to derive the “average ACR phenotype” (20).

To better utilise the longitudinal data values, we derived a novel phenotype that models the trajectory of covariate-adjusted ACR measurements over time. For this, we used Bayesian multilevel modelling to fit average slopes as the trajectories. We opted to use a multilevel modelling approach as this allowed us to borrow information across subjects, in particular to improve parameter estimation for subjects with few data points available (and thus an uncertain trajectory). Explicitly, we used the brm function within the brms R package (26) which utilises the Stan software to fit the following Bayesian linear multivariate multilevel model:

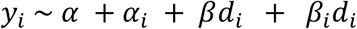

where *y*_*i*_ is the residual for subject *i* measured at *d*_*i*_ years since their diagnosis. Parameters *α* and *β* represent the fixed effects and have independent improper flat priors, whilst parameters *α*_*i*_ and *β*_*i*_ represent random effects and are sampled from a multivariate normal prior with zero mean and an unknown variance-covariance matrix. We used the default prior for this variance-covariance matrix (see https://cran.r-project.org/web/packages/brms/vignettes/brms_overview.pdf for more details) and default parameter values, but increased the iterations per chain from 2000 to 8000 in order to ensure sufficient mixing of chains. We extracted the mean posterior slope coefficient for each subject (comprising both the fixed and random coefficients; *β* + *β*_*i*_), and used these subject-specific values as our “trajectory phenotypes”.

### GWAS Analysis

For study samples with both genotype and phenotype data available (*N* = *1584*), we used the Gemma software (27) to perform genetic association tests for each of the 5 imputed data sets for each of our 3 phenotypes (“latest ACR phenotype”, “average ACR phenotype” and “trajectory phenotype”). The genomic control lambda values ranged between 0.95 and 0.99.

Because ACR has a right-skewed distribution, there is a potential for exaggerated p-values. To ensure our inference was robust, we repeated the complete analysis of each phenotype 1000 times with permuted phenotypes, and of a standard normal phenotype. For each null analysis, we saved the minimum p-value, and used the relationship between the minimum p-values from permuted and standard normal phenotype analysis to interpolate a more conservative adjusted p-value, upon which we based our GWAS inference.

We used the conventional significance threshold of *p* < *5* × *10*^−*8*^ to call genome-wide significant signals and *p* < *1* × *10*^−*6*^ for suggestive signals. To identify index SNPs in associated regions we selected the SNP with the smallest p-value in each 200-kb region. To aid comparison across phenotypes, we report standardised estimated effect sizes (*β*) and standard errors (SEs) (standardised so that the variance of the phenotype values equal 1).

### Fine-Mapping Association Signals

We used Bayesian fine-mapping to prioritise the likely causal variants in associated regions, assuming a single causal variant per-region (25, 26). To define loci for fine-mapping we included all variants 200-kb either side of the index-SNP, and used the corrcoverage R package (29) to derive posterior probabilities of causality (PPs) for each SNP in the region, and to generate 95% credible sets.

To ensure that the single causal variant assumption made by corrcoverage was robust, we also performed fine-mapping using SuSiE (27, 28), which allows for multiple causal variants, using reference LD data from European samples in the 1000 Genomes Project (32).

### Replication Analysis

We used the Type 1 Diabetes Knowledge Portal (T1DKP; https://t1d.hugeamp.org/, accessed 10 Feb 2022) to scrutinise significant signals identified by our analysis. We used results from the PheWAS association analysis for studies conducted on samples of European ancestry. In practice, the relevant studies on T1DKP were for the SUMMIT consortium (30, 31). We recorded the sample size, effect size (odds ratio due to case-control GWAS) and p-value for the relevant SNPs.

### Data and Resource Availability

GWAS summary statistics will be made publicly available on the GWAS Catalog. Code to perform the analysis is available via https://github.com/annahutch/T1D-ACR and https://github.com/chr1swallace/acr-p-values. Phenotype data is available upon request.

## Results

### Novel Trajectory Phenotype

We describe a novel “trajectory phenotype” which effectively models the slope of covariate-adjusted ACR measurements over time. For this, we use a multilevel modelling approach that allows information to be borrowed across individuals, thereby protecting against extreme phenotype values for individuals with few data points. In practice, if an individual has many data points and therefore a more complete trajectory, then the method attenuates the slope of the observed trajectory only slightly towards the mean slope (Figs. 1A, 1B). In contrast, if an individual has very few data points and therefore a more uncertain trajectory, then the method more heavily attenuates the slope towards the mean (Figs. 1C, 1D).

**Fig. 1:**
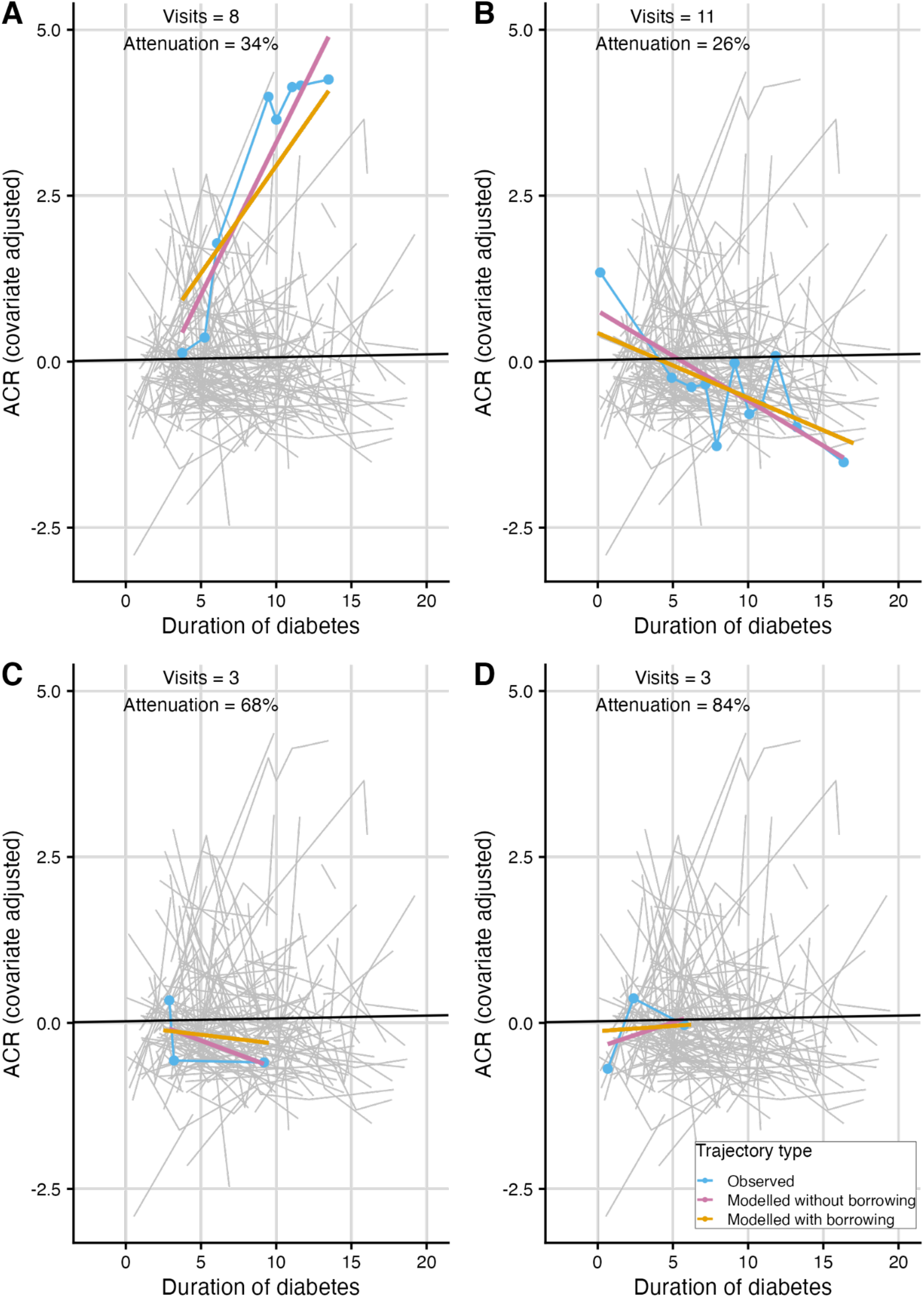
Figure to illustrate modelling ACR trajectories to derive the trajectory phenotype. In plots of covariate-adjusted ACR measurements against duration of diabetes, we show the observed trajectory (line between data points for each individual), the modelled trajectory without borrowing information across individuals (straight “best fit” line fitted to data points for each individual) and the modelled trajectory with borrowing across individuals (from which the slope equals the “trajectory phenotype”). Panels (A) and (B) highlight trajectories for individuals with many data points available, where the borrowing only attenuates the slope of the trajectory slightly. Panels (C) and (D) highlight trajectories for individuals with only three data points available, where the borrowing attenuates the slope of the trajectory more heavily. The grey lines represent a random sample of observed trajectories. The average trajectory across all subjects is shown in black. The number of visits and the attenuation between the trajectory modelled with borrowing and without borrowing is given in each panel. Attenuation was calculated as *1* − *slope of trajectory with borrowing*/ *slope of trajectory without borrowing*.

### Comparison with Related Phenotypes

Previous studies of ACR have used cross-sectional ACR measurements (8, 11–13) or average ACR values (20). In our data, these values were highly correlated (*r* = *0*.*77*). Our trajectory phenotype was also highly correlated with the latest ACR values (*r* = *0*.*66*) and moderately correlated with the average ACR values (*r* = *0*.*49*). This indicates that an individual’s trajectory is related to their average ACR, with more rapidly progressing trajectories corresponding to higher ACR values at the last visit.

Some understanding of the differences between these phenotypes may be revealed by considering individuals identified as having extreme phenotypic values. We found that the average ACR phenotype tended to identify those individuals with a single (or small number of) extreme ACR measurements (Fig 2). In contrast, the trajectory phenotype highlights those individuals with strongly increasing (or decreasing) ACR measurements over time, thus monitoring albumin progression. The latest ACR phenotype only uses information from the most recent visit and is agnostic to any ACR measurements taken before this time.

**Fig. 2:**
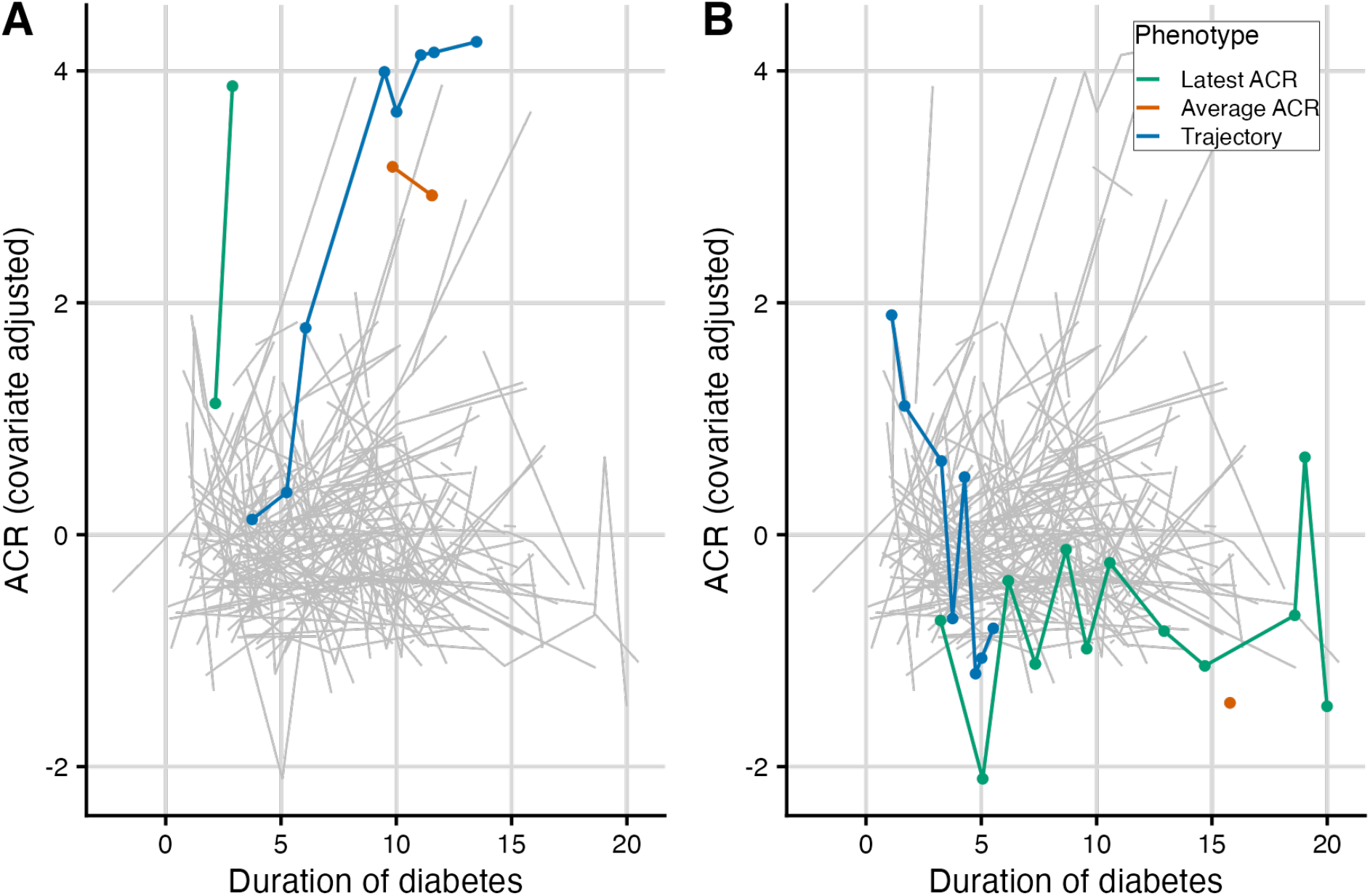
Figure to illustrate observed trajectories for individuals with the highest and lowest phenotype values. In plots of covariate-adjusted ACR measurements against duration of diabetes, we highlight the observed trajectories for individuals with the (A) highest and (B) lowest phenotype value for each of the three phenotypes. The grey lines show a random sample of observed trajectories.

### GWAS Results

We performed GWAS for all three ACR phenotypes (Fig. 3, Supplementary Fig. 8). At a suggestive significance threshold of *p* < *1* × *10*^−*6*^, two signals were found to associate with the trajectory phenotype, three with the latest ACR phenotype (including one of the trajectory associated SNPs) and one with the average ACR phenotype (Table 2).

**Table 2:** SNPs with suggestive evidence of association (*p* < *1* × *10*^−*6*^) in any of the three GWAS phenotypes. For each index SNP, the table contains the rsID, the chromosome, the base position (bp, hg19), the alleles (reference > effect), the estimated effect size (beta), the standard error of the estimated effect size (SE), the observed and adjusted p-values and the closest protein coding gene. Note that rs145715205 appears twice, once for the trajectory and once for the latest phenotype.

| rsID | Chr | bp | Alleles | Beta | SE | P raw | P adj | Closest gene |
| --- | --- | --- | --- | --- | --- | --- | --- | --- |

**Table 2:** SNPs with suggestive evidence of association ( $p < 1 \times 10^{-6}$ ) in any of the three GWAS phenotypes.
| *Trajectory* |  |  |  |  |  |  |  |  |
| --- | --- | --- | --- | --- | --- | --- | --- | --- |
| rs115207889 | 1 | 64897735 | T>G | 0.929 | 0.171 | $6.53 \times 10^{-8}$ | $9.97 \times 10^{-7}$ | *CACHD1* |
| **rs145715205** | **5** | **20409042** | **A>C** | **0.883** | **0.135** | $8.9 \times 10^{-11}$ | **$2.61 \times 10^{-8}$** | ***CDH18*** |
| *Average ACR* |  |  |  |  |  |  |  |  |
| rs112200619 | 1 | 65861210 | T>C | 0.743 | 0.141 | $2.20 \times 10^{-7}$ | $6.07 \times 10^{-7}$ | *LEPROT* |
| *Latest ACR* |  |  |  |  |  |  |  |  |
| **rs150766792** | **4** | **17193426**<br>**8** | **A>G** | **0.729** | **0.125** | $6.94 \times 10^{-9}$ | **$4.04 \times 10^{-8}$** | ***GALNTL6*** |
| **rs145715205** | **5** | **20409042** | **A>C** | **0.668** | **0.134** | $6.37 \times 10^{-9}$ | **$1.66 \times 10^{-8}$** | ***CDH18*** |
| rs187626285 | 7 | 73817796 | G>A | 0.828 | 0.158 | $1.81 \times 10^{-7}$ | $5.89 \times 10^{-7}$ | *GTF2IRD1* |
| rs61947176 | 13 | 37885033 | G>A | 0.774 | 0.146 | $1.41 \times 10^{-7}$ | $4.85 \times 10^{-7}$ | *CSNK1A1L* |

**Fig. 3:**
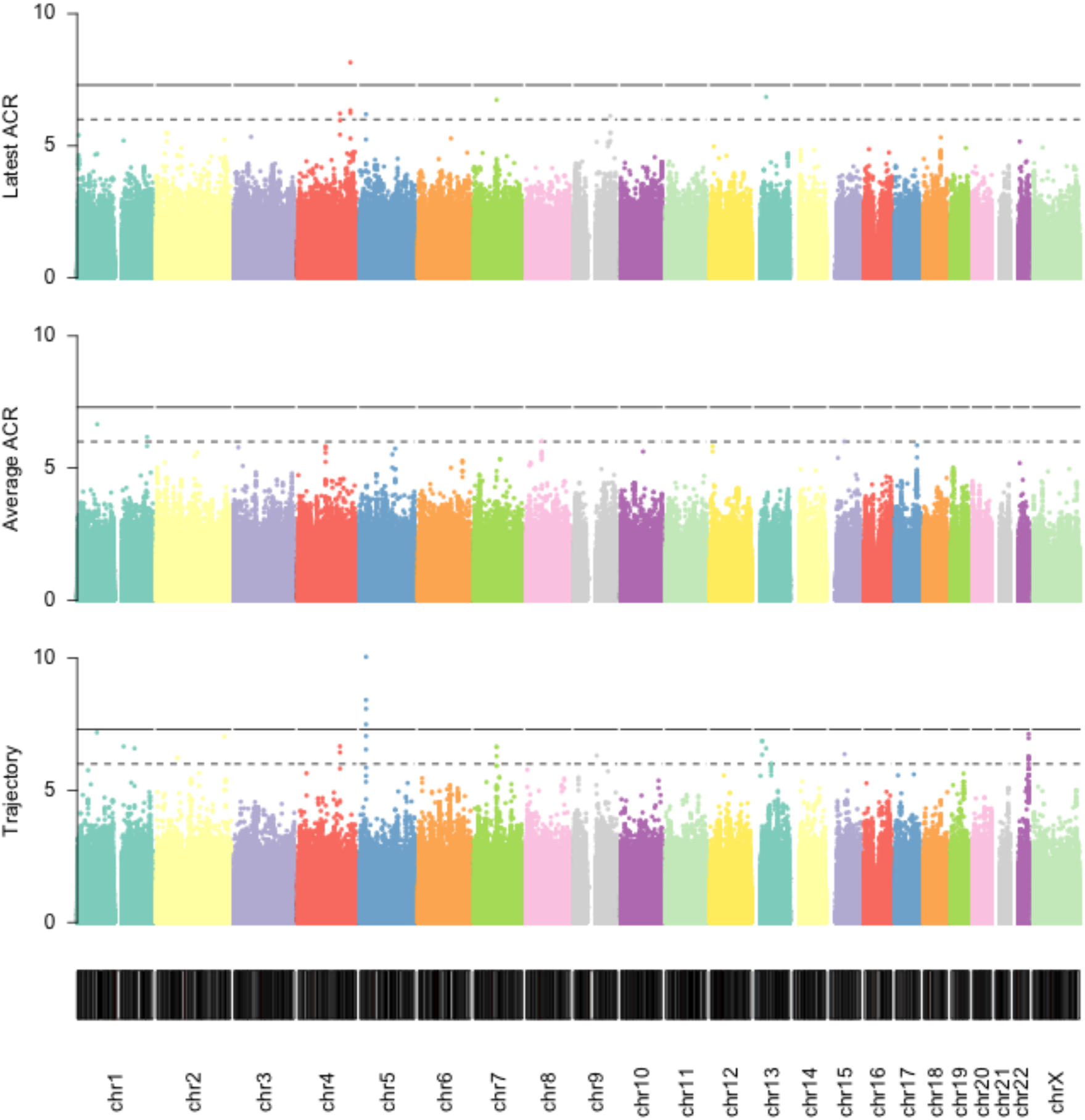
Manhattan plots of GWAS results for (A) latest ACR phenotype (B) average ACR phenotype and (C) trajectory phenotype. Solid line on Manhattan plots show genome-wide significance threshold (*p* = *5* × *10*^−*8*^) and dashed line shows suggestive significance threshold (*p* = *1* × *10*^−*6*^). The y-axis is the −*log*_*10*_ transformed p-value and the x-axis is the genomic position.

Of these suggestive signals, two exceeded the more stringent genome-wide significance threshold of *p* < *5* × *10*^−*8*^. The first of these is for the latest ACR phenotype, and has index SNP rs150766792 for which the alternative allele (G) is relatively rare in European populations (*MAF* = *0*.*0219*) (35). The SNP rs150766792 is closest to the long intergenic non-coding gene, *LINC02431* (27-kb from the start of the gene). The closest protein coding gene to rs150766792 is *GALNTL6* (800-kb to TSS).

The second genome-wide significant signal is for the trajectory phenotype and is driven by genetic variants residing in the first intron of the *CDH18* gene on chromosome 5. The alternative allele (C) at the index SNP, rs145715205, is relatively rare in European populations (*MAF* = *0*.*0139*) (35). Interestingly, the latest ACR also showed suggestive association with rs145715205 in *CDH18* (*p* = *6*.4 × 10^−7^) and also with rs79607490 (*P* = *6*.*0* × *10*^−*7*^), which is upstream of *PCDH18* on chromosome 4, another member of the cadherin superfamily.

### Examining Significant Signals

We used Bayesian fine-mapping to prioritise the likely causal variants in the two genomic regions that reached genome-wide significance. In the *GALNTL6* gene region the index SNP, rs150766792, was found to be the most likely causal variant (*PP* = *0*.*89*) and the 95% credible set contained 2 additional variants (rs140921123 *PP* = *0*.*040*; rs12502900 *PP* = *0*.*054*) (Fig. 4, Supplementary Fig. 9).

**Fig. 4:**
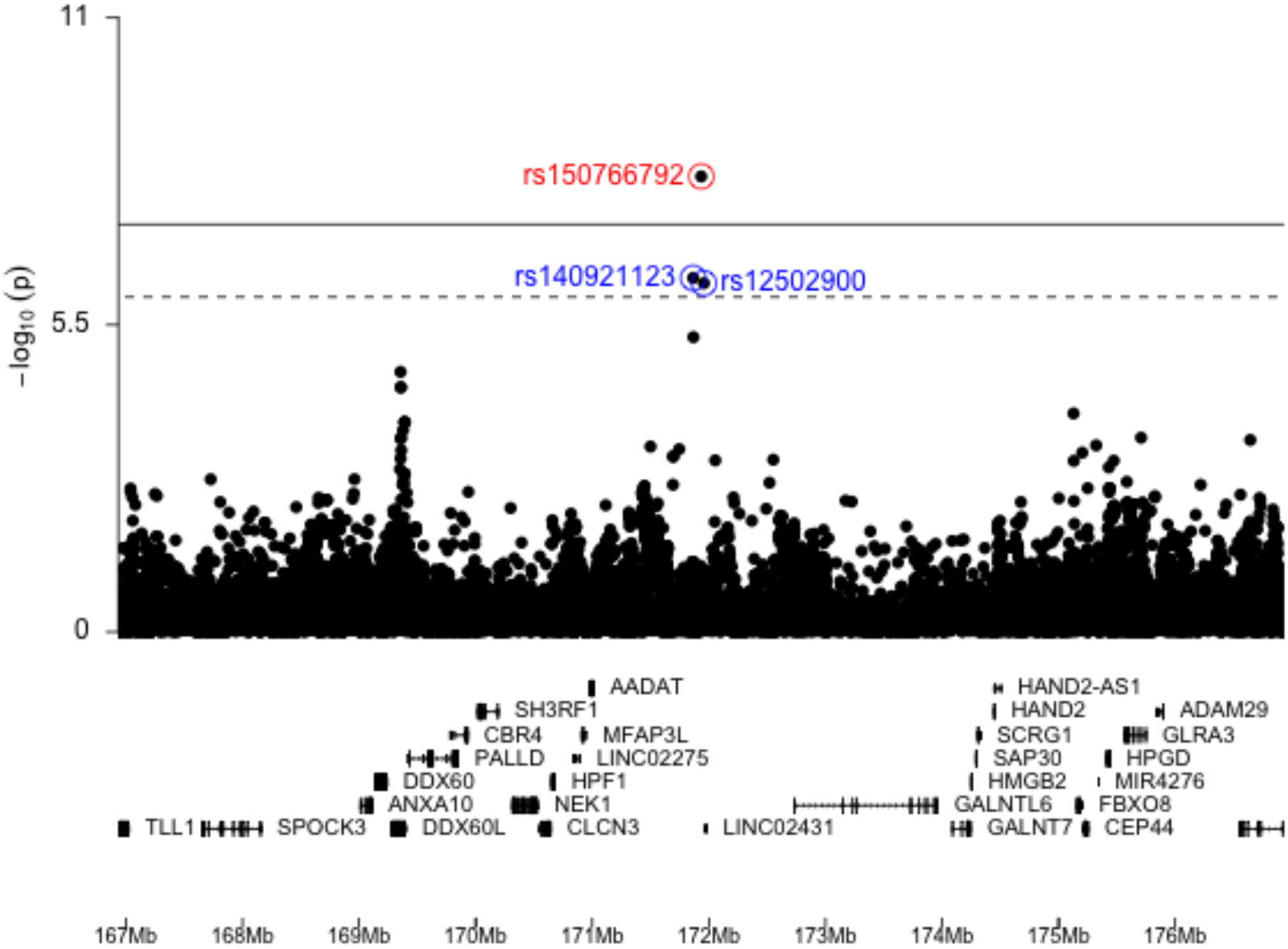
Manhattan plot for the *GALNTL6* gene region (chr54:167000000-176000000). The index SNP in the region is highlighted and labelled in red. The two other SNPs included in the 95% credible set are highlighted and labelled in blue. Genes and genomic position (hg19) along chromosome 4 are shown in the bottom panel.

In a PheWAS association analysis on T1DKP, we found that the most strongly associated phenotypes with rs150766792 were microalbuminuria and all diabetic kidney disease, from the SUMMIT consortium (Table 3). Reassuringly, the effect sizes were directionally consistent with those from our GWAS, whereby the effect allele (G) increased risk. The index SNP rs150766792 is correlated with the two other SNPs in the credible set (*r*^*2*^ = *0*.*4346* with rs140921123 and *r*^*2*^ = *0*.*5695* with rs12502900; estimated using 1000 Genomes Project European populations using the LDmatrix tool in LDlink (32,36)), and these SNPs also associated with microalbuminuria and all diabetic kidney disease in the SUMMIT consortium on T1DKP.

**Table 3:** Replication results for rs150766792 from the Type 1 Diabetes Knowledge Portal (T1DKP; https://t1d.hugeamp.org/) in studies where subjects were of European ancestry. The effect allele for rs150766792 was G and the reference allele was A.

| Study | Sample size | OR | P-value | Reference |
| --- | --- | --- | --- | --- |
| Microalbumin in subjects with T1D or T2D | 5,474 | 1.70 | 0.0003718 | (30 ,31) |
| Microalbumin in subjects with T2D | 3,464 | 1.90 | 0.001107 | (34) |
| Microalbumin in subjects with T1D | 1,747 | 1.41 | 0.1077 | (33) |
| All diabetic kidney disease in subjects with T1D or T2D | 9,307 | 1.50 | 0.001 | (30 ,31) |
| All diabetic kidney disease in subjects with T2D | 4,414 | 1.79 | 0.001436 | (34) |
| All diabetic kidney disease in subjects with T1D | 4,599 | 1.24 | 0.1554 | (33) |

In the *CDH18* gene region the index SNP, rs145715205, was found to be the most likely causal variant (*PP* = *0*.*975*) (Fig. 5). The PheWAS on T1DKP identified urinary sodium excretion as the most strongly associated phenotype with rs145715205 (*p* = *0*.*007*) (37), with urinary albumin-to-creatinine ratio the second most strongly associated phenotype (*p* = *0*.*019*) (35). SNP rs145715205 was not present in the SUMMIT data set so we were unable to check replication in this cohort.

**Fig. 5:**
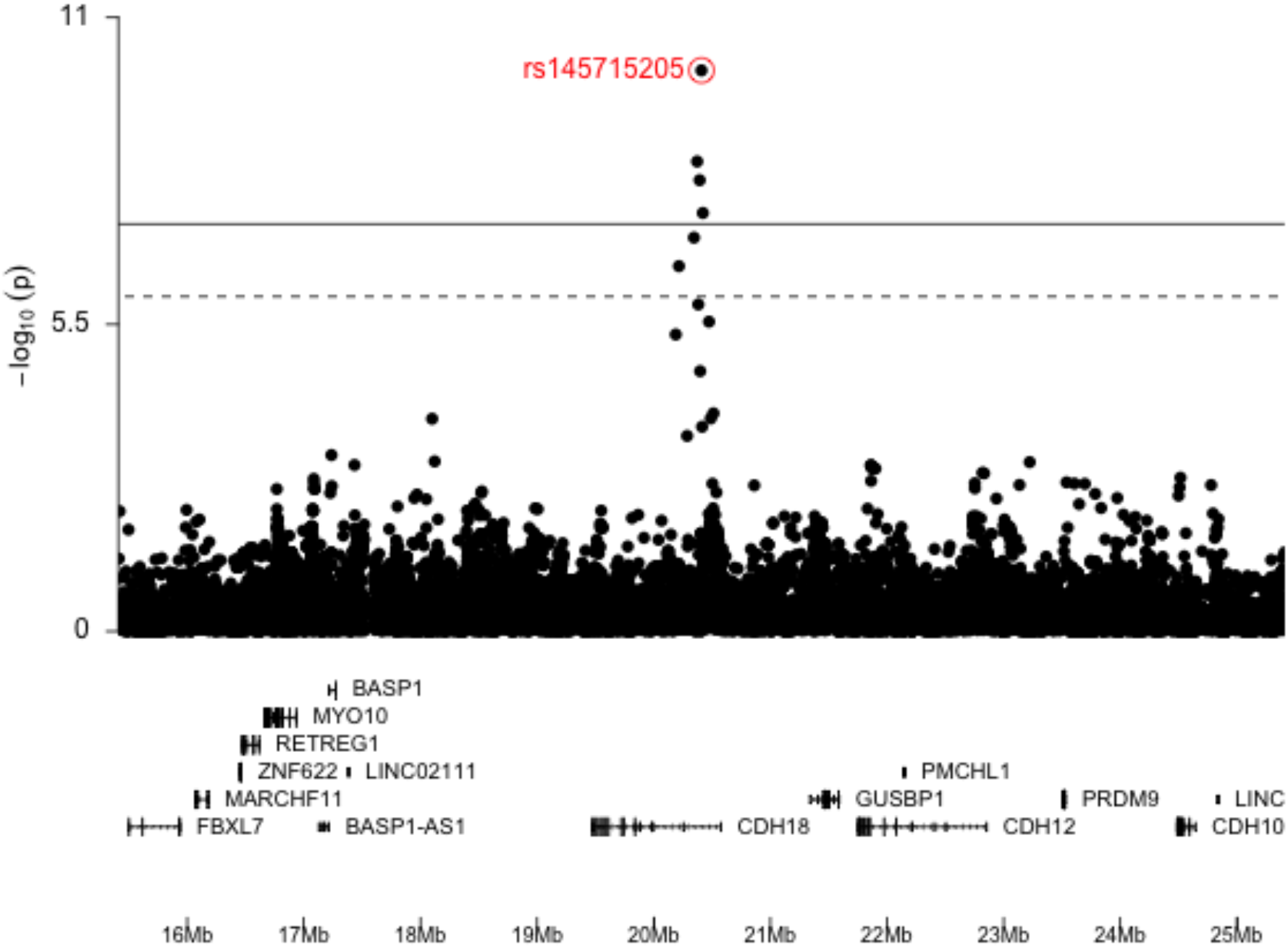
Manhattan plot for the *CDH18* gene region (chr5:16000000-25000000). The index SNP in the region is highlighted and labelled in red. Genes and genomic position along chromosome 5 are shown in the bottom panel (hg19).

## Discussion

We have described a novel albumin excretion phenotype, the trajectory phenotype, that is based on longitudinal data. This complements albumin-related phenotype definitions used in previous studies which are typically based on cross-sectional data by quantifying the slope, and not simply the central or final location of ACR measures. Estimated slopes will have a much higher degree of uncertainty for individuals with only a few measurements compared to those with more measurements. We accounted for this by taking a multilevel approach which naturally attenuates the most uncertain slopes towards the global mean, and reduces the risk of outlying observations having undue influence on GWAS estimates. Given the variability in numbers of observations per individual, we considered this approach was likely to be more robust than other approaches for summarising longitudinal data, such as latent trajectory modelling that groups individuals into trajectory groups whereby individuals within the same trajectory group have similar trajectories. Whilst such approaches have the advantage of being able to accommodate non-linear trajectories, this requires many more degrees of freedom, which in turn requires more observations, amplifying the difficulty assigning accurate phenotypes to individuals with fewer observations.

To the best of our knowledge, no other study has derived a phenotype that models longitudinal ACR changes for use as a continuous trait in GWAS. Whilst we used this phenotype to identify genetic variants which predict better or worse ACR trajectories, a similar approach could be considered for studying potential non-genetic predictors. Such approaches could help towards the development of a personalised prediction tool to help guide therapeutic decisions, for example commencing statin usage earlier if genetic or other risk factors are present.

Our signal close to *GALNTL6* (lead SNP rs150766792) for the latest ACR phenotype was replicated in the SUMMIT cohort, which was reassuring because our cross-sectional latest ACR phenotype more closely resembled the microalbuminuria and DKD case definitions in the SUMMIT GWAS than the other phenotypes. We identified a novel signal in the *CDH18* gene (lead SNP rs145715205) when using our trajectory phenotype. We were unable to replicate this signal, which may reflect that no previous studies have looked for genetic associations with albuminuria progression in type 1 diabetes patients.

The *CDH18* gene, located in the region of chromosome 5 encodes for cadherin 18, which belongs to the large cadherin superfamily, a class of calcium-dependent trans-membrane proteins. Cadherin 18 is expressed in various tissues but appears to be confined primarily to the central nervous system (38). SNPs within *CDH18* have been previously associated with gestational diabetes (39) and with metabolic syndrome-related traits, such as obesity, insulin resistance and blood pressure (40). Although there is no clear evidence yet for *CDH18* to be expressed in the kidney or vascular system, and there is no previous evidence of association with albumin excretions or more general DKD, the role in metabolic syndrome-related traits is of interest. Indeed, urinary albumin excretion and DKD are closely associated with features of the metabolic syndrome, such as obesity, insulin resistance and hypertension (41,42).

The *GALNTL6* gene encodes the membrane-bound protein N-acetylgalactosaminyltransferase like 6, which has a significant role in the pathway of protein glycosylation (43,44). There are no previous reports of a role of this gene in the context of diabetes and its vascular complications. However, a potential role could be speculated given that protein glycosylation is a well known process implicated in the pathogenesis of vascular complications of diabetes (45).

It will therefore be important to validate our findings, for example in larger study cohorts or cohorts of differing ancestry. Functional studies will also be required to investigate the functional roles of rs145715205 and rs150766792 in albuminuria and its progression in patients with type 1 diabetes. Of interest, this study also identified a new approach to assess longitudinal ACR data, which could be applied not only in future GWAS studies, but could also help towards the development of a personalised prediction tool to help guide therapeutic decisions, for example commencing more intensive drug interventions earlier if genetic or other risk factors are present.

## Data Availability

All data produced in the present study are available upon reasonable request to the authors

## Acknowledgments

The authors gratefully acknowledge the contribution made to this study by David Dunger who was the Chief Investigator of the ORPS/NFS and AdDIT studies prior to his death in July 2021. We would like to thank Rany Salem for advice on genotype data cleaning. Data on diabetic kidney disease has been contributed by the SUMMIT Consortium and has been downloaded from mccarthy.well.ox.ac.uk (http://mccarthy.well.ox.ac.uk/publications/2018/SUMMIT_DKD/) on 9/02/2022.

## Author Contributions

A.H performed the analysis and wrote the manuscript. M.L.M. conceived the hypothesis, prepared data, contributed to discussion and reviewed/edited manuscript. C.W helped conceive the analysis pipeline, supervised the analysis, contributed to discussion and reviewed/edited manuscript. P.B, F.J.C, S.T.C, J.J.C, R.N.D, D.D, E.A.D, J.E.D, K.C.D, T.W.J, F.H.M, S.M.M, A.N contributed to data collection and critical review of the manuscript. W-M.C, S.O-G, S.S.R contributed to genetic analyses.

