## Supplemental Material for "Genome-wide association study of longitudinal urinary albumin excretion in patients with type 1 diabetes"

#### GWAS Genotyping, Quality Control, and Imputation

The genotype data was received in three genotyping groups, and for convenience we refer to these groups as group 1, group 2 and group 3. Quality control (QC) and imputation were performed separately within each group and there was no sample overlap between groups.

##### Groups 1 and 2

The data for samples in groups 1 and 2 had already been quality controlled (Supplementary Table 1). As described by Salem et al. 2019[1] , the study samples underwent genotyping and QC centrally at the University of Virginia. Samples were genotyped on the HumanCore BeadChip array (Illumina, San Diego, CA), which contains approximately 250,000 genome-wide tag SNPs and 200,000 exome-focused variants. All samples were passed through a stringent QC protocol, including recalling with zCall (a calling algorithm specifically designed for rare SNPs from arrays), aligning variants to build hg19, updating variant names using 1000 Genomes as a reference and filtering low-quality variants (e.g., call rates ,95% and excessive deviation from Hardy–Weinberg equilibrium) and samples (e.g., call rates ,98%, sex mismatch, extreme heterozygosity). We included an additional QC step to remove 7 SNPs in group 1 and 244 SNPs in group 2 which were flagged by PLINK (version 1.9)[2] as heterozygous in males but did not reside in the pseudo-autosomal region of chromosome X.

| | **Group 1** | | **Group 2** | |
|---|---|---|---|---|
| | **Samples** | **SNPs** | **Samples** | **SNPs** |
| **Initial raw genotype files** | **200** | **542,585** | **1363** | **547,644** |
| Remove SNPs from combined QC step | | -12,192 | | -13,975 |
| Remove subjects from combined QC step | -3 | | -8 | |
| Remove SNPs that couldn't be mapped to hg19 | | -12,732 | | -12,537 |

| | | | | |
|---|---|---|---|---|
| Remove SNPs from study-specific QC step | | -222,025 | | -162,261 |
| Remove subjects from study-specific QC step | -2 | | -32 | |
| Remove non-European samples | -3 | | -129 | |
| Remove problematic heterozygous haploid SNPs | | -17 | | -244 |
| **Final QC'ed data** | **192** | **296,233** | **1194** | **359,529** |

Supplementary Table 1: Table describing the pre-imputation QC steps implemented for samples in genotyping groups 1 and 2. “Combined QC" identifies SNPs for removal with call rates less than 99%, those residing on the Y chromosome in female samples, those with Mendel inconsistencies, those with concordant rates less than 99%, those with Y call rates less than 99% and those in Hardy-Weinberg disequilibrium. In addition, “Combined QC " identifies samples for removal which are mislabeled as male or female and those missing more than 5% of genotypes. “Study specific QC " identifies monomorphic SNPs and those in Hardy-Weinberg disequilibrium. In addition, “Study specific QC" identifies samples with extreme heterozygosity values. These QC steps had already been implemented, and are described in [1]. We included an additional SNP QC step to remove any problematic heterozygous SNPs.

We used the Michigan Imputation Server[3] to impute missing genotypes for samples in groups 1 and 2, using the Haplotype Reference Consortium (HRC) panel of 64,976 human haplotypes at 39,235,157 SNPs as the reference data (version r1.1 2016)[4]. To prepare the data for imputation we used the HRC-1000G-check-bim.pl script written by Will Rayner (https://www.well.ox.ac.uk/~wrayner/tools/), which updates the strands, alleles and genomic coordinates of the variants to reflect HRC/hg19, and removes variants with large differences in allele frequencies compared to HRC ($> 20\%$) or ambiguous alleles (AT/GC variants with $MAF > 40\%$). This step removed 3180 variants in group 1 and 13365 variants in group 2.

We used the VcfCooker tool (https://genome.sph.umich.edu/wiki/VcfCooker) to convert the data to per-chromosome bgzipped VCF files for input into the imputation server, and specified that the samples were of European ancestry and that any variants with low imputation quality ($R^2 < 0.3$) should be discarded. We ran the imputation job on the server in the "QC and imputation mode'" with phasing using Eagle (version 2.4)[5]. After completion, we downloaded the imputed data and used the ic.pl script written by Will Rayner (https://www.well.ox.ac.uk/~wrayner/tools/) to check the quality of the results. The post-imputation QC reports are publicly available (group 1: https://github.com/annahutch/T1D-GWAS/blob/main/Group1.html, group 2: https://github.com/annahutch/T1D-GWAS/blob/main/Group2.html) and show that the imputed data is of high quality.

Following imputation we removed samples with extreme proportions of homozygous variants, defined as having homozygosity proportions $< 0.828$ or $> 0.834$ in group 1 (5 samples removed; Supplementary Fig. 1A) and $> 0.888$ in group 2 (3 samples removed; Supplementary Fig. 2A), since low proportions may indicate low sample quality whilst high values may indicate inbreeding. We also removed 321,570 monomorphic SNPs in group 1 (Supplementary Fig. 1B) and 532,889 monomorphic SNPs in group 2 (Supplementary Fig. 2B).

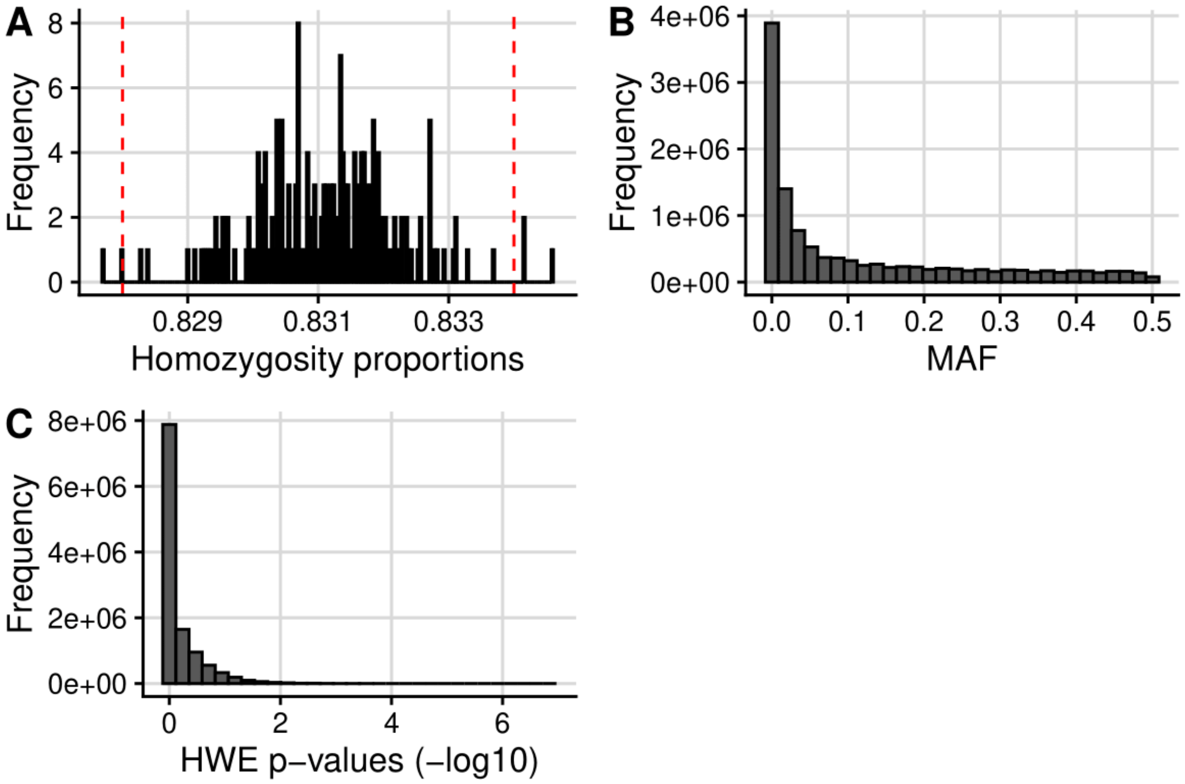


Supplementary Fig. 1: Plots to facilitate post-imputation quality control for study samples in genotyping group 1. (A) Histogram of per-sample homozygosity proportions with red dashed lines showing the thresholds used to remove samples due to extreme homozygosity proportions (<0.828 or >0.834). (B) Histogram of MAF values. (C) Histogram of HWE p-values.

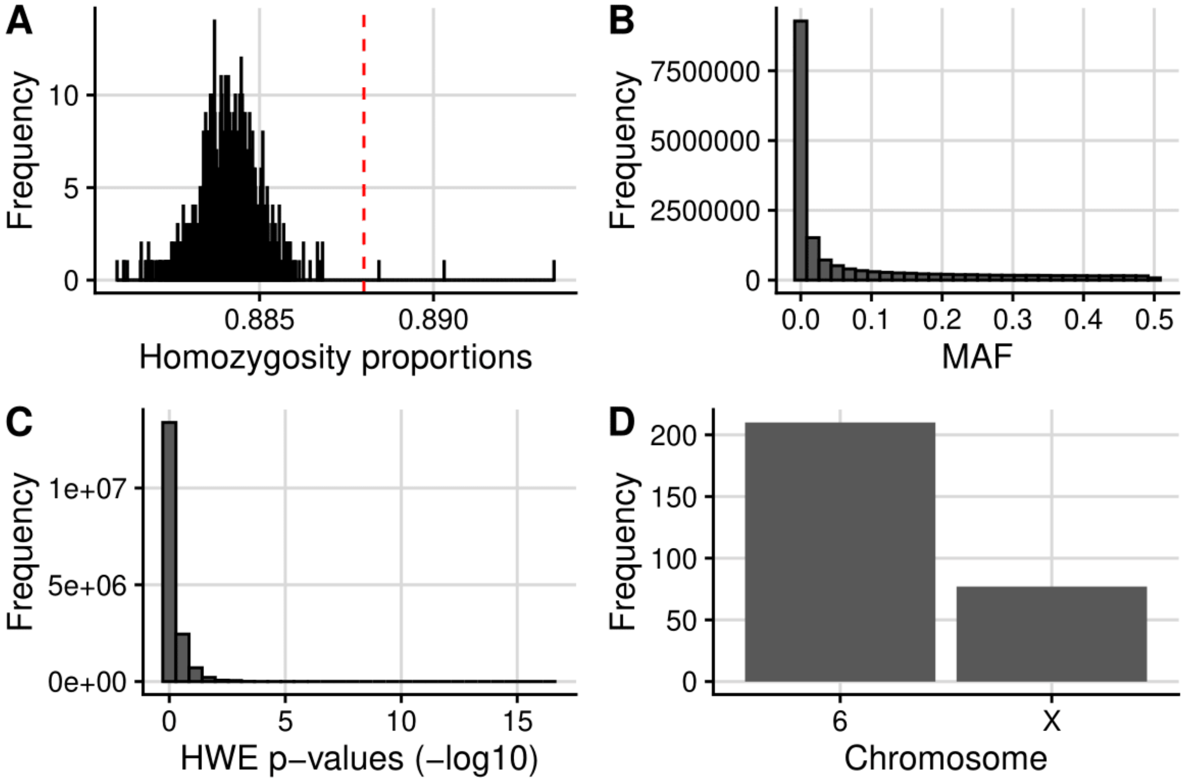


Supplementary Fig. 2: Plots to facilitate post-imputation quality control for study samples in genotyping group 2. (A) Histogram of per-sample homozygosity proportions with a red dashed line showing the threshold used to remove samples due to extreme homozygosity proportions (>0.888). (B) Histogram of MAF values. (C) Histogram of HWE p-values. (D) The number of SNPs with HWE $p < 1e - 10$ that resided on each chromosome (SNPs with HWE $p < 1e - 10$ were only found on chromosomes 6 or X).

Deviations from Hardy-Weinberg equilibrium (HWE) are likely to be the consequence of genotyping error, inbreeding or population stratification[6]. Since the samples in groups 1 and 2 with evidence of inbreeding (quantified by high homozygosity proportions) had already been removed, and those of non-European ancestry had also been removed (Supplementary Table 1), it was likely that any deviations from HWE at this stage of the analysis were due to genotyping errors. Generally, departure from HWE is examined in control samples because SNPs that strongly associate with the trait of interest are not expected to be in HWE in case samples. However, all samples in groups 1 and 2 were cases, meaning that no control data was available. In group 1, the minimum HWE p-value was

$1.412e-07$ (Supplementary Fig. 1C), whilst for group 2 the minimum HWE p-value was $3.935e-17$ (Supplementary Fig. 2C). All of the 287 SNPs in group 2 with HWE $p < 1e-10$ resided on either chromosome 6 or the X chromosome (Supplementary Fig. 2D). This was reassuring because it is known that the MHC strongly associates with T1D, and so the SNPs with evidence of departure from HWE on chromosome 6 are likely reflecting the strong association of the MHC with T1D. Moreover, PLINK only uses female samples to evaluate departure from HWE for SNPs residing on the X chromosome (since male samples are hemizygous), which may be problematic due to a decrease in power (when the sample sizes of groups 1 and 2 were already small) and for additional reasons which are discussed in [7] .

The imputed and quality controlled data consisted of 11,483,371 SNPs in 187 samples (group 1) and 16,730,386 SNPs in 1191 samples (group 2).

##### Group 3

We downloaded PLINK formatted genotype data for 966 samples in genotyping group 3 and removed any samples with incorrectly formatted sample IDs (13 with “rep**" and 7 with “unq**"). For the 69 remaining samples with replicates within this genotyping group (due to being sequenced on different wells or plates), we selected the replicate with the best SNP coverage. We then converted the data from GRCh38 to hg19 coordinates and discarded 2059 SNPs which could not be converted between the builds. The data had not yet been quality controlled and so we implemented a pre-imputation QC pipeline to ensure that the study data used for imputation was of high quality (Supplementary Table 2).

| | **Group 3** | |
|---|---|---|
| | **Samples** | **SNPs** |
| **Initial raw genotype file** | **966** | **745,577** |

| | | |
|---|---|---|
| Remove samples with incorrectly formatted sample IDs | -20 | |
| Remove duplicate samples | -69 | |
| Remove SNPs that could not be converted from GRCh38 to hg19 | | -2059 |
| Remove samples with imputed sex not matching reported sex | -2 | |
| Remove samples with high missingness | -6 | |
| Remove samples with high proportions of homozygous variants | -3 | |
| Remove SNPs with high missingness | | -36293 |
| Remove SNPs with low HWE p-value | | -209 |
| Remove monomorphic SNPs | | -68,397 |
| Remove non-European samples | -35 | |
| **Final QC'ed data** | **841** | **638,619** |

Supplementary Table 2: Table describing the pre-imputation QC steps implemented for samples in genotyping group 3.

Firstly, we removed two samples incorrectly listed as females (Supplementary Fig. 3A) and used the inferred sex for 556 samples with missing sex information. We then removed any samples with either $> 3\%$ of SNPs missing or with extreme proportions of homozygous variants ($> 0.83$) (Supplementary Fig. 3B). We checked for any related individuals in the genotyping group by first pruning the variants so that no pair of SNPs within 50-kb were correlated ($r^2 > 0.2$) (using a step size of 5) and then using the --genome flag in PLINK to derive pairwise IBD proportions (Supplementary Fig. 3C). We made a list of the related samples but did not remove these at this stage of the analysis, since related samples do not affect imputation results. We also removed SNPs missing in more than 2% of individuals (Supplementary Fig. 3D).

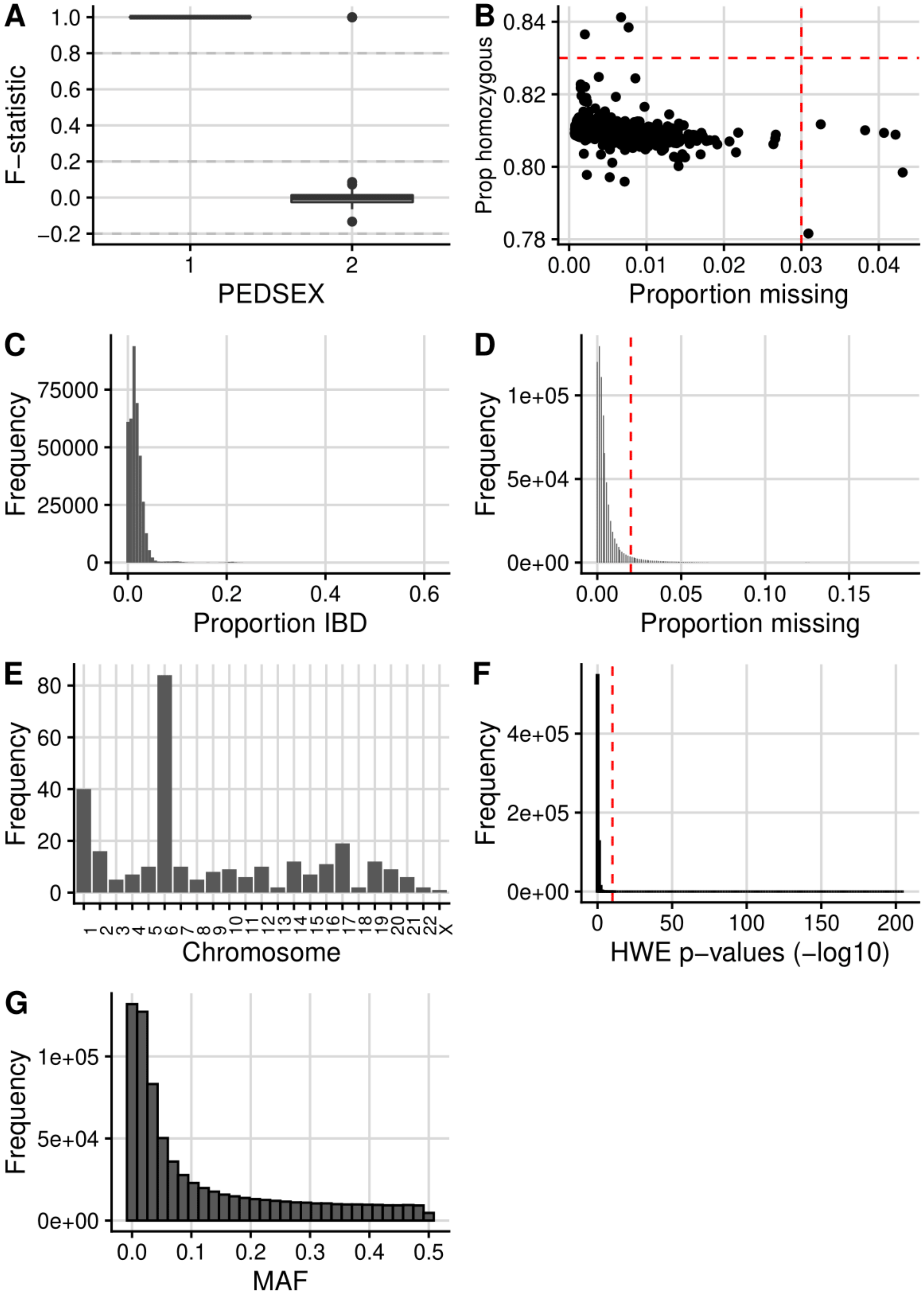

A
F−statistic
PEDSEX
B
Prop homozygous
Proportion missing
C
Frequency
Proportion IBD
D
Frequency
Proportion missing
E
Frequency
Chromosome
F
Frequency
HWE p−values (−log10)
G
Frequency
MAF

Supplementary Fig. 3: Plots to facilitate pre-imputation QC for study samples in genotyping group 3. (A) Box plot showing the F-statistic (actual X chromosome homozygosity estimate) values for samples labelled as male (PEDSEX=1) and female (PEDSEX=2). A male call is made if F>0.8 and a female call is made if F<0.2 (grey dashed lines). (B) Per-sample homozygosity proportions against the proportions of SNPs missing in each sample, with red dashed lines showing the thresholds used to remove samples (proportion homozygosity >0.8 or proportion missing >0.03). (C) Histogram of proportion IBD values (P(IBD=2) + 0.5*P(IBD=1)) for each pair of samples used to determine related pairs of individuals. (D) Histogram of per-SNP missingness with a red dashed line at proportion missing equal to 0.02. (E) The number of SNPs with HWE $p < 1e - 10$ on each chromosome. (F) Histogram of HWE p-values for SNPs not residing on chromosome 6 or in the *PTPN22* gene region (chr6:114256433-114514381) with a red dashed line at HWE $p = 1e - 10$. (G) Histogram of MAF values.

We examined the SNPs with small HWE p-values and found that most SNPs with HWE $p < 1e - 10$ were on chromosome 6 or chromosome 1 (Supplementary Fig. 3E). This is likely due to the fact that all of the samples were cases and that there is a strong association between the MHC (chromosome 6) and *PTPN22* (chromosome 1) with T1D (although it was interesting that chromosome 11 didn't contain more variants with evidence of departure from HWE, due to the strong association of *INS* and T1D). We removed 209 SNPs with HWE $p < 1e - 10$ which did not reside on chromosome 6 or in the *PTPN22* gene region (chr6:114,256,433-114,514,381, which covers the *PTPN22* gene plus 100-kb either side) (Supplementary Fig. 3F). We also removed 68,397 monomorphic SNPs (Supplementary Fig. 3G).

We next checked for any non-European samples using the 1000 Genomes data as reference. Following the "Ancestry estimation based on reference samples of known ethnicities" vignette (available from https://cran.r-project.org/web/packages/plinkQC/vignettes/AncestryCheck.pdf) from the plinkQC R package, we firstly pruned the study data to exclude known regions of high LD and so that no pair of

SNPs with 50-kb were correlated ($r^2 > 0.2$) (using a step size of 5). We then filtered the reference data to only contain the pruned SNPs in the study data set and then matched SNPs between the reference data and the study data based on genomic position and alleles. We merged the two data sets and performed PCA using the --pca flag in PLINK. We then used the plinkQC::evaluate_check_ancestry function to estimate the ancestry of the study samples from the PCA results. Briefly, the function uses principal components 1 and 2 to find the centre of the known European reference samples, and any study samples whose Euclidean distance from the centre falls outside the radius specified by the maximum Euclidean distance of the reference samples multiplied by the chosen europeanTh value (default = 1.5) are labelled as non-European. This analysis identified 35 samples as non-European and we excluded these from the analysis (Supplementary Fig. 4).

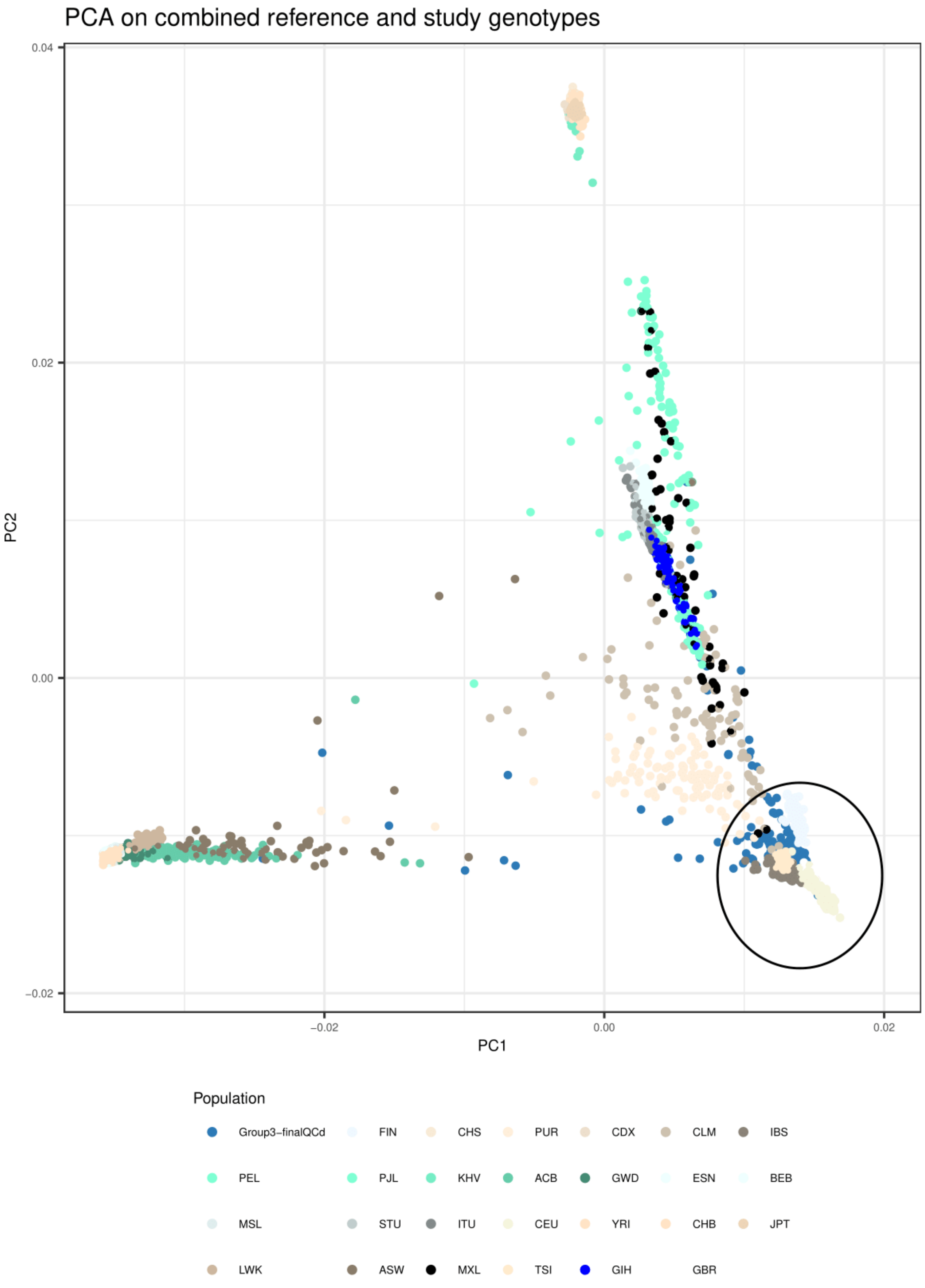


Supplementary Fig. 4: Figure generated using the plinkQC::evaluate_check_ancestry function to identify any samples of non-European ancestry, using the 1000 Genomes data as reference.

As before, we used Will Rayner's HRC-1000G-check-bim.pl script to prepare the data for imputation, which removed 18473 SNPs with large differences in allele frequencies compared to HRC (>20%) or ambiguous alleles (AT/GC variants with MAF>40%). We then used the Michigan Imputation Server with the HRC reference panel for imputation. The post-imputation QC report generated using Will Rayner's ic.pl script is publicly available (group 3: https://github.com/annahutch/T1D-GWAS/blob/main/Group3.html) and shows that the imputed data is of high quality.

Following imputation, we removed 7 samples with extreme proportions of homozygous variants, defined as having homozygosity proportion $< 0.877$ or $> 0.885$ (Supplementary Fig. 5A) and removed 657,771 monomorphic SNPs (Supplementary Fig. 5B). For the 302 SNPs with HWE $p < 1e - 10$, the vast majority resided on chromosome 6 and the X chromosome as expected, but 6 resided on chromosome 10 and eight resided on chromosome 18 (Supplementary Fig. 5C, Supplementary Fig. 5D). Upon further investigation, the minimum HWE p-values for SNPs residing on chromosome 10 was $3.491e - 12$ and the minimum HWE p-values for SNPs residing on chromosome 18 was $3.372e - 26$. We subsequently removed these SNPs.

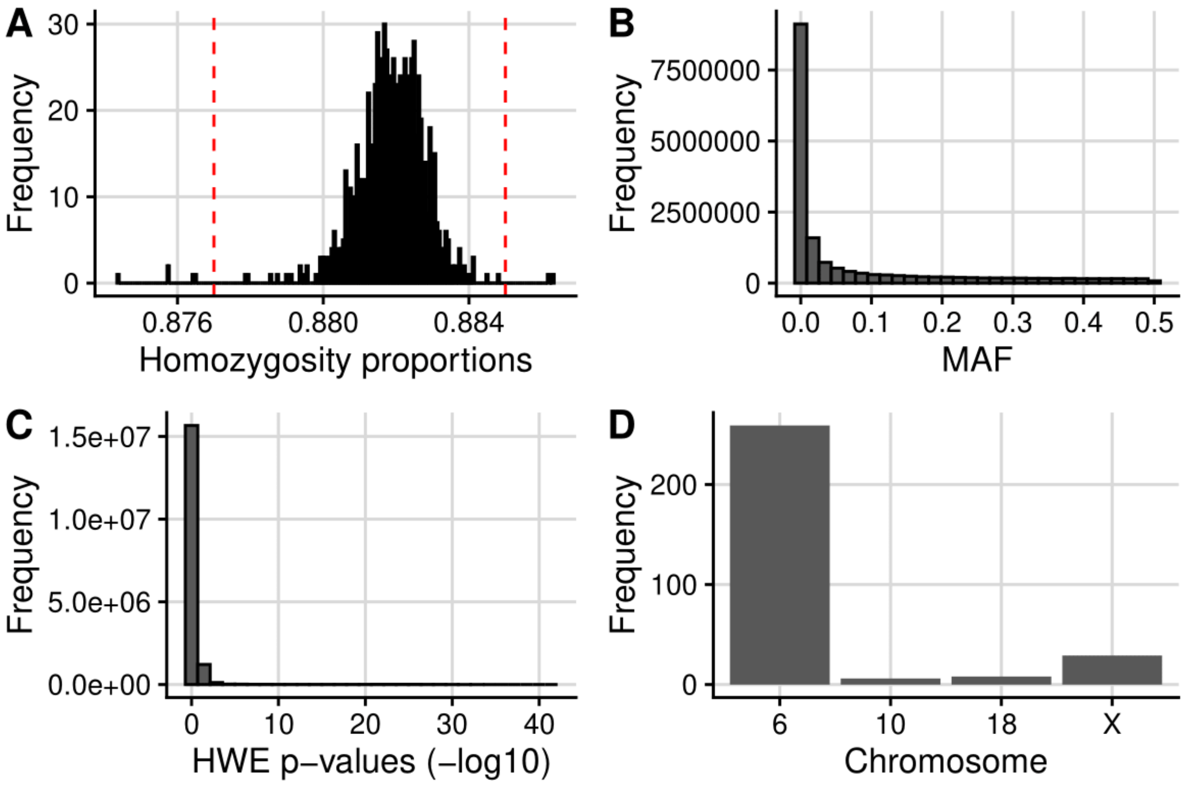


Supplementary Fig. 5: Plots to facilitate post-imputation quality control for samples in genotyping group 3. (A) Histogram of per-sample homozygosity proportions with a red dashed line showing the threshold used to remove samples due to extreme homozygosity proportions (<0.877 or >0.885). (B) Histogram of MAF values. (C) Histogram of HWE p-values. (D) The number of SNPs with HWE p<1e-10 that resided on each chromosome (SNPs with HWE $p < 1e - 10$ were only found on chromosomes 6, 10, 18 or X).

The imputed and quality controlled data for group 3 consisted of 16,583,186 SNPs in 827 samples.

#### Rubin's rules

In this section we describe the derivation of pooled effect sizes, pooled standard errors and pooled p-values using Rubin's rules [8] (https://bookdown.org/mwheymans/bookmi/rubins-rules.html).

The pooled effect size is the geometric mean of the effect size calculated from each of the imputed data sets. For SNP j

$$\beta_{pooled,j} = \sum_{i=1}^{m} \beta_{i,j}$$

where $m$ is the number of imputed data sets ($m = 5$).

The pooled standard error for SNP j is

$$SE_{pooled,j} = \sqrt{V_{Total,j}}$$

where

$$V_{Total,j} = V_{W,j} + V_{B,j} + \frac{V_{B,j}}{m}$$

and $V_{W,j}$ is the within imputation variance for SNP j:

$$V_{W,j} = \frac{1}{m}\sum_{i=1}^{m} SE_{i,j}^{\ 2}$$

and $V_{B,j}$ is the between imputation variance for SNP j:

$$V_{B,j} = \frac{\sum_{i=1}^{m} (\beta_{i,j} - \beta_{pooled,j})^2}{m-1}.$$

To calculate a p-value from the pooled effect size and standard error, we need to define the degrees of freedom. For this we use the adjusted degrees of freedom described by Barnard and Rubin (1999):

$$df_{Adjusted} = \frac{df_{old} \times df_{observed}}{df_{old} + df_{observed}}$$

where

$$df_{old} = \frac{m-1}{\lambda^2},$$

and

$$df_{observed} = \frac{(n-k)+1}{(n-k)+3} \times (n - k)(1 - \lambda).$$

$n$ is the sample size of the imputed dataset, $k$ is the number of parameters to fit and lambda is:

$$\lambda = \frac{V_B + \frac{V_B}{m}}{V_{Total}}.$$

We then obtain a two-sided p-value using the t distribution.

### Supplementary Figures

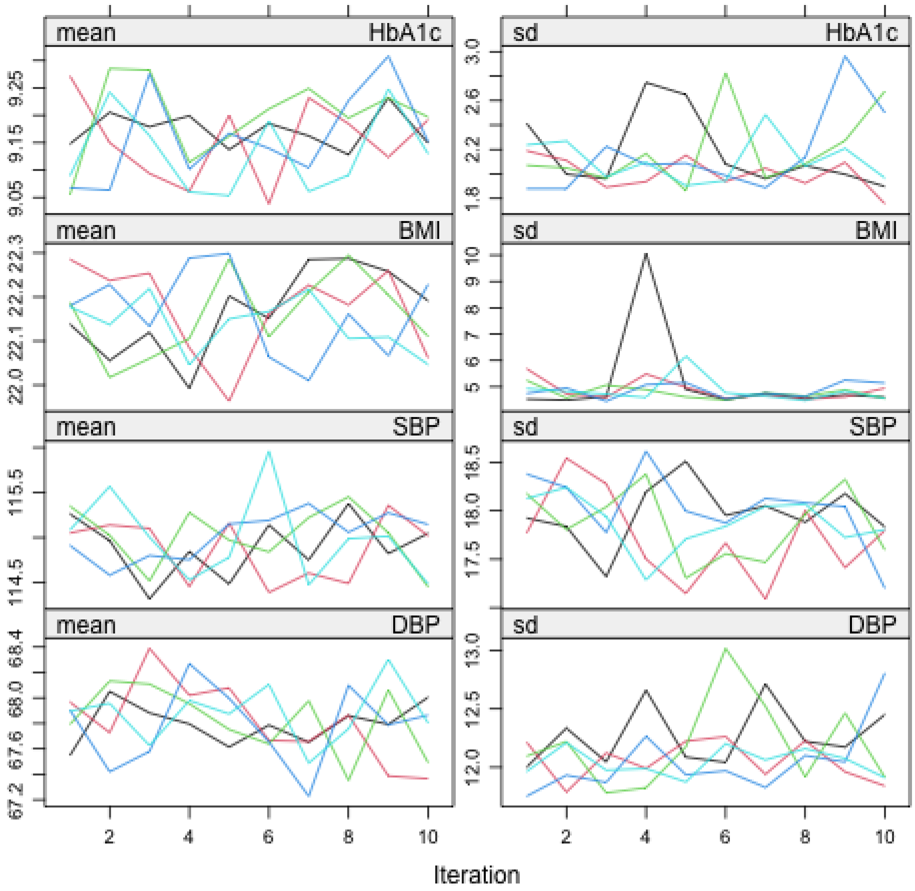


Supplementary Fig. 6: Trace plots for MCMC draws from the multiple imputation. The mean (left) and standard deviation (right) of the imputed values against the iteration number for each of the 5 replications.

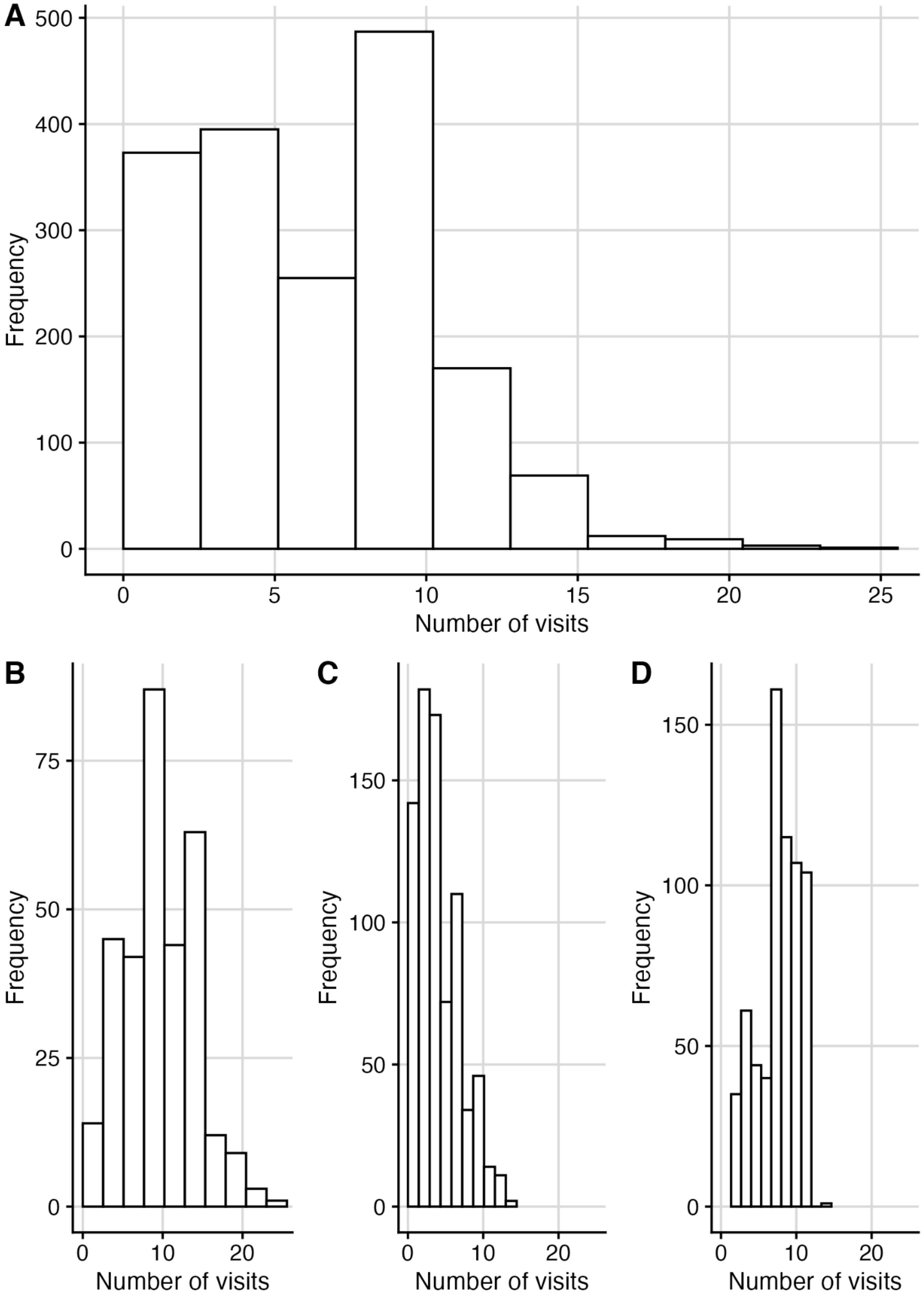
A
B
C
D
Frequency
Number of visits
0
100
200
300
400
500
5
10
15
20
25
50
75
150

Supplementary Fig. 7: Histograms showing number of visits per individual. Panel (A) is for all individuals across all three cohorts, (B) is for individuals in ORPS (C) is for individuals in NFS and (D) is for individuals in AdDIT.

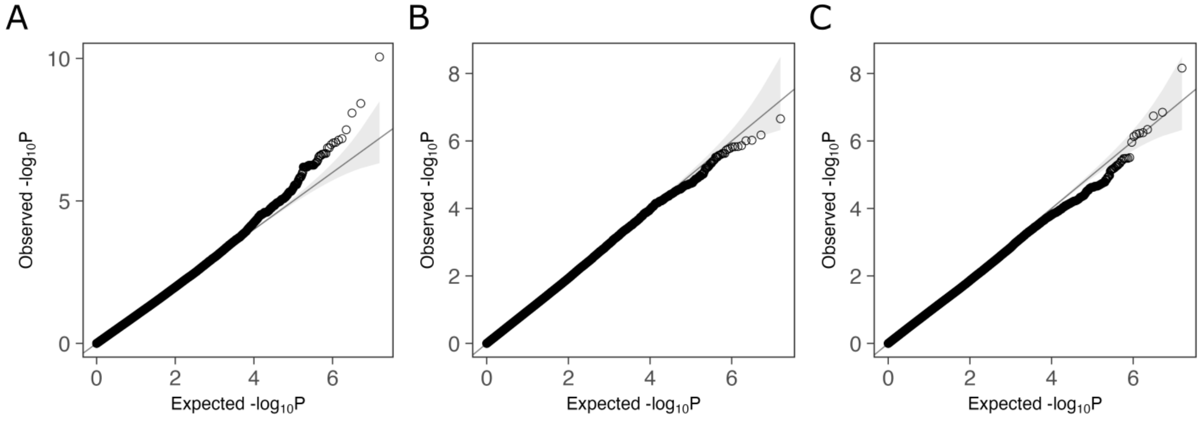


Supplementary Fig. 8: QQ plots of GWAS results for analysis of (A) trajectory phenotype (B) average ACR phenotype and (C) latest ACR phenotype.

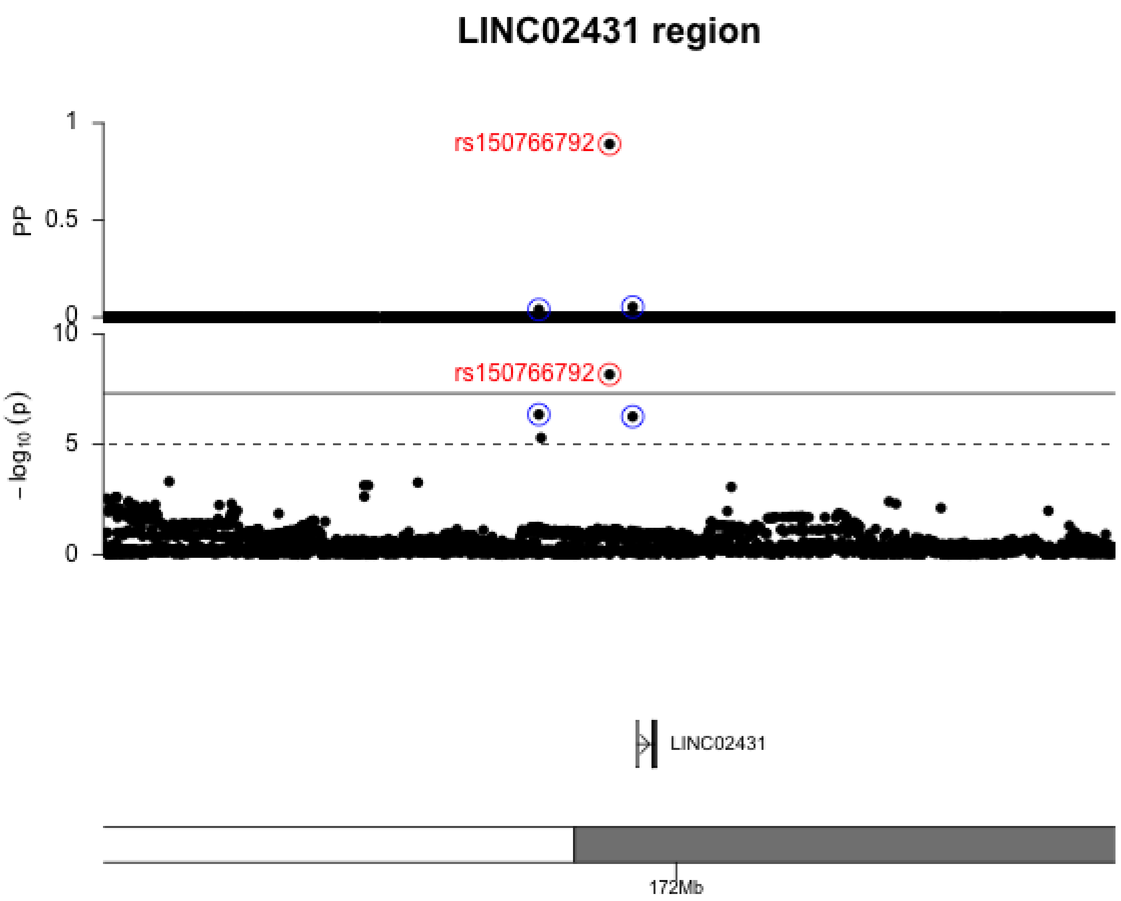


Supplementary Figure 9: Fine-mapping and GWAS results in the *LINC02431/ GALNTL6* gene region (chr4:171434268-172434268). First panel shows posterior probabilities (PPs) from Bayesian fine-mapping and the second panel shows -log10 transformed p-values from GWAS. The index SNP in the region is highlighted and labelled in red. The two other SNPs included in the 95% credible set (rs140921123 and rs12502900) are highlighted in blue. Genes and genomic position along chromosome 4 are shown in the bottom panel.
